# Exploring athlete pain assessment experiences and priorities; A two-part qualitative series of athlete and physiotherapist interactions. Part One. “Gauging and discerning” - Athlete & physiotherapist pain assessment experiences and interactions

**DOI:** 10.1101/2024.01.05.24300908

**Authors:** Ciarán Purcell, Caoimhe Barry Walsh, Garett Van Oirschot, Brona M Fullen, Tomás Ward, Brian M Caulfield

**Affiliations:** School of Public Health, Physiotherapy and Sports Science, University College Dublin, Dublin, Ireland; Insight SFI Research Centre for Data Analytics, Dublin, Ireland; School of Allied Health, University of Limerick, Limerick, Ireland; Physical Activity for Health Research Cluster, University of Limerick, Limerick, Ireland; Sports and Human Performance Centre, University of Limerick, Limerick, Ireland; Ageing Research Centre, University of Limerick, Limerick, Ireland

**Keywords:** ‘Pain Measurement’, ‘Musculoskeletal Pain’, ‘Athletic Injuries’, ‘Athletic Performance’, ‘Focus Groups’, ‘Pain Management’

## Abstract

**Objectives:** To explore the content (subjective questions, objective tools and outcome measures) and discuss the nature (qualitative elements and wider considerations) of the athlete pain assessment by facilitating shared understandings of athlete and sports physiotherapists.

**Design:** Qualitative Research using a hermeneutic phenomenological approach.

**Methods:** We carried out focus groups comprising a deliberate criterion sample using a constructivist perspective. We developed a topic guide and used reflexive thematic analysis. We developed codes, candidate themes and finalised themes iteratively, and employed a critical friend to add depth to our analysis. Our paper follows the consolidated criteria for reporting qualitative research (COREQ) guidelines.

**Results:** We completed five focus groups comprising twelve athletes (five female, seven male) and four sports physiotherapists (four male). Three final themes (and eight subthemes) were created; (i) Measures, Scales and Dimensions (value and limitations of tools and scales, multidimensional methods, making sense and interpreting), (ii) Connect, Listen and Learn (the pain interview and athlete’s story, forging the athlete-clinician connection), (iii) Lighthouse in the Storm: providing direction for athletes in pain. (information overload and indecision, a beacon of direction; the role of the physiotherapist, the burden of expectation; challenges for physiotherapists)

**Conclusion:** We described and explored the phenomena of pain assessment in sport including current pain assessment strategies. Comprehensive multidimensional assessment methods that preserve the athlete-clinician therapeutic relationship and facilitate optimal communication are priorities for future research and practice.

## Introduction

A comprehensive biopsychosocial pain assessment that includes affective (emotional), cognitive (understandings and appraisals) and socioenvironmental (sport, home, work/school, social, cultural and environmental) as well as the more frequently used neurophysiological (pain characteristics and qualities) and biomechanical local and wider biomechanical considerations) aspects has been proposed to capture the full extent of an athlete’s pain experience. ^1^ ^2^ Understanding the impact of these aspects on an athlete’s sport and wider life facilitates the planning and delivery of more effective management strategies.^3^ In a recent scoping review, we identified gaps in pain assessment practice specifically around the affective, cognitive and socioenvironmental aspects of pain assessment.^4^ Contemporary models outline the impact of wider aspects such as stress, sports environment, support networks and coping strategies on athlete pain experience and injury prevention.^5^ ^6^ Clear and effective communication allows athletes to develop relationships with their support network and to access coping strategies they may not be able to otherwise. This is in line with contemporary research that highlights the importance of effective communication between all members of the athlete support staff in injury prevention.^7^ In this series we explore one of the key relationships and communication opportunities in an athlete’s pain journey, the athlete-physiotherapist pain assessment. To date, little has been published exploring athlete pain assessment, and to the best of our knowledge, this is the first study to combine both athlete and physiotherapist experiences and perspectives. Our objectives were to explore the content (subjective questions, objective tools and outcome measures) and discuss the nature (qualitative elements and wider considerations) of an assessment for athletes with upper and lower limb pain

## Methods

We conducted a qualitative study with mixed focus groups that combined athletes and sports physiotherapists and followed the consolidated criteria for reporting qualitative research (COREQ) guidelines.^8^ Ethical permission was granted for our study by the UCD Human Research Ethics Committee (LS-22-40-Purcell-Caulfield).

### Positionality statement and team background

We viewed perceptions of athlete upper and lower limb pain and pain assessment from a constructivist perspective taking a pragmatic approach and drawing on the relevant knowledge from elements of empirico-analytical, interpretive and critical research paradigms.^9^ As the primary interviewer I (CP) recognise that being a sports physiotherapist and PhD athlete pain researcher influences the dynamic of discussions, I am unavoidably enmeshed, and I embrace this to understand athlete and clinician upper and lower limb pain experiences with a reflexive approach. ^9^ ^10^. GvO PhD candidate and sports physiotherapist with seventeen years of experience acted as moderator for the first two focus groups to ensure all voices were heard and took field notes of the setting and context and IOC Athlete Pain domains discussed. ^1^ BC a Professor in physiotherapy with twenty years of research experience acted as a neutral observer for the first session. CBW, Physiotherapist and PhD researcher in the field of chronic conditions with experience in the conduct of focus groups, acted as a critical friend for the qualitative analysis.

### Participants

We recruited a deliberate criterion-based sample of: (i) athletes (of varying age, gender, sport, and competition level) through university and local sports clubs and (ii) sports physiotherapists (with a minimum of three years of postgraduate experience working with athletes as part of their weekly caseload from a variety of sports and clinical settings) through the Irish Society of Chartered Physiotherapists in Sports and Exercise Medicine group via email and other communication channels and networks. All athletes had been assessed for pain in the upper or lower limb in the past year, and all sports physiotherapists had assessed such patients also. To facilitate engagement we offered participants their preference of face-to-face and online (zoom) focus group settings (which were run separately). We used a reflexive sampling strategy with data collection and analysis following each focus group informing the total sample size. We ceased sampling and recruitment when sufficient depth of information on athlete pain experience and athlete-physiotherapist pain assessment experience was gathered rather than seeking objective data saturation in line with our active constructivist approach. ^11^

### Protocol

We sent participants a pre-participation questionnaire to capture demographic information including their sports background and previous pain and injury history. We asked physiotherapists about their experience working in sports, additional qualifications and any current or prior sports participation. We developed a topic guide (Appendix A) using an established step-by-step approach; brainstorming, ordering, timing and phrasing of questions, obtaining feedback from peers, revising and piloting.^12^ The topic guide moved from broad questions concerning athlete pain understandings and experiences to reflections on current pain assessment practice before identifying priorities for an athlete upper and lower limb pain assessment framework. We altered the sequencing of questions slightly in subsequent focus groups in line with an iterative and reflexive approach.^10^ We allocated two hours for the sessions to ensure enough time for all components. We set ground rules establishing confidentiality, equity and respect for diverse views and opinions before each group. We moderated the power imbalance between physiotherapists and athletes by emphasising the importance of the athlete’s voice asking for athletes’ opinions to be shared before clinicians. Where possible we ensured athletes and physiotherapists who previously worked together were separated. Our initial successful pilot focus group of two athletes was followed up with four focus groups combining athletes and physiotherapists in line with methodological recommendations to facilitate interaction through dialogue and capture various perspectives in a shared space.^13^

### Analysis

We completed the transcription verbatim and transcripts were checked against the audio recordings for accuracy. The full, uncoded and anonymised set of transcripts has been published in an online data repository (10.17632/t47tw94mzd). We chose reflexive thematic analysis as it was congruent with both our data collection and epistemological position.^10^ Following initial observations and notes CP carried out initial coding of the entire transcripts (both semantic and latent). During the data analysis process, we assigned athletes alphanumeric participant IDs beginning with the letter “A” and physiotherapists alphanumeric IDs beginning with ‘’P”. Athletes and physiotherapists were talking about and experiencing the same concepts so we merged the data for coding and analysis. Participant IDs were known only to CP, the lead researcher. Once participant IDs were allocated and analysis was complete all records of participant information were deleted ensuring anonymity Athlete and physiotherapist data were merged for coding and analysis. Subsequent analysis was completed to identify codes that were present in (i) athletes only, (ii) physiotherapists only and (iii) codes that were present in both athletes and physiotherapists. One-third of the transcripts were given to CBW to code independently and we discussed, compared and refined initial codes using a critical friend approach to incorporate breadth and variation in experiences and perspectives.^14^ We updated coding where necessary, with similar codes being highlighted and grouped to form rough clusters of candidate themes. The team reviewed candidate themes in the context of the research question with consideration given to the data available to support each theme. We merged some themes and adjusted others at this refinement stage. We developed an initial thematic map of the refined themes before we applied titles and created a final thematic map. (Appendix B) In this paper, Part One of this series, we report on data primarily from questions focused on athlete pain assessment experience.

## Results and Discussion

Participant demographics are summarised in Table 1. All final themes exploring athlete pain assessment included a mixture of codes from all three categories (i, ii and iii in the data analysis section above). Figure 1 displays the finalised themes and codes. Figure 2 is a thematic map of the themes and subthemes relevant to athlete pain assessment.

**Figure 1.**
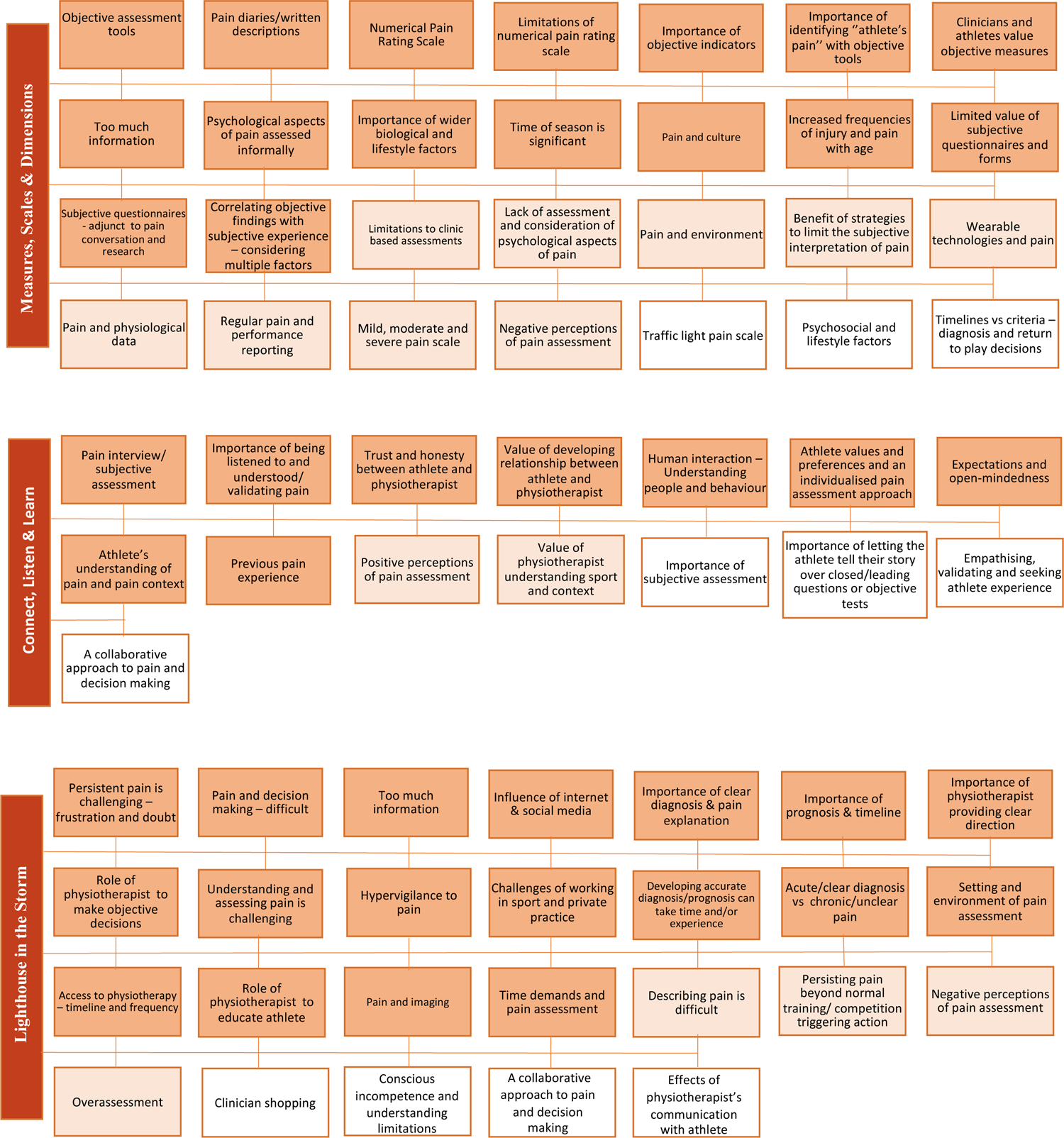
Themes (Measures, Scales and Dimensions; Connect, Listen and Learn; Lighthouse in the Storm: providing direction for athletes in pain) and codes. Dark shading – indicates codes that were present in athletes and physiotherapists. Light shading – indicates codes that were present in athletes only, No shading – indicates codes that were present in physiotherapists only

**Figure 2.**
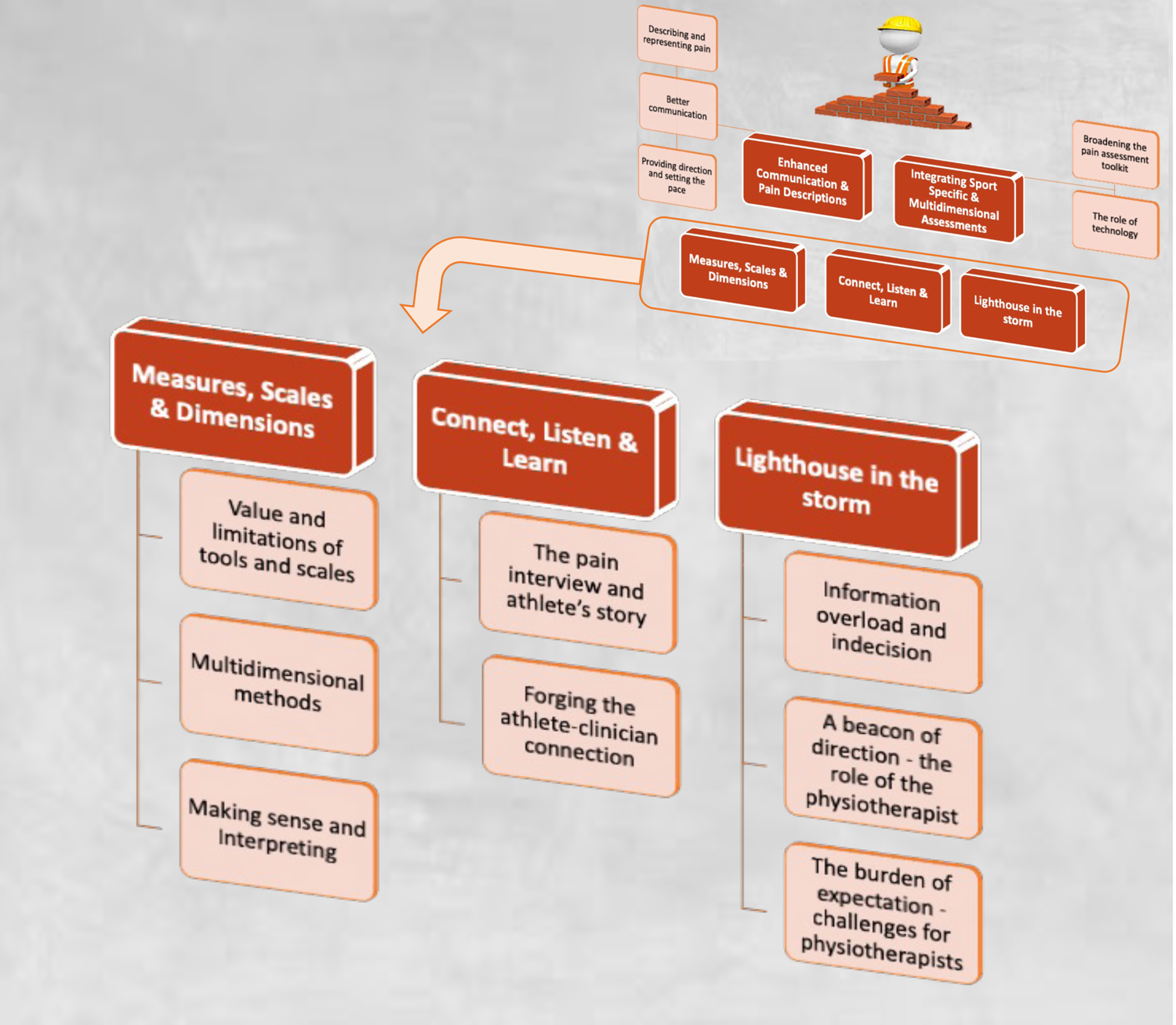
Athlete pain assessment experience themes and subthemes. The themes for each part of this series represent a row in the priorities for the pain assessment pyramid, this paper presents the bottom or foundation row of the pyramid. This comprises the three themes that describe the “athlete pain assessment” experience alongside the constituent subthemes. In the top row, the “priorities and directions for athlete pain assessment” themes and subthemes will be presented in Part Two and will therefore build on the themes from this paper.

**Table 1.**
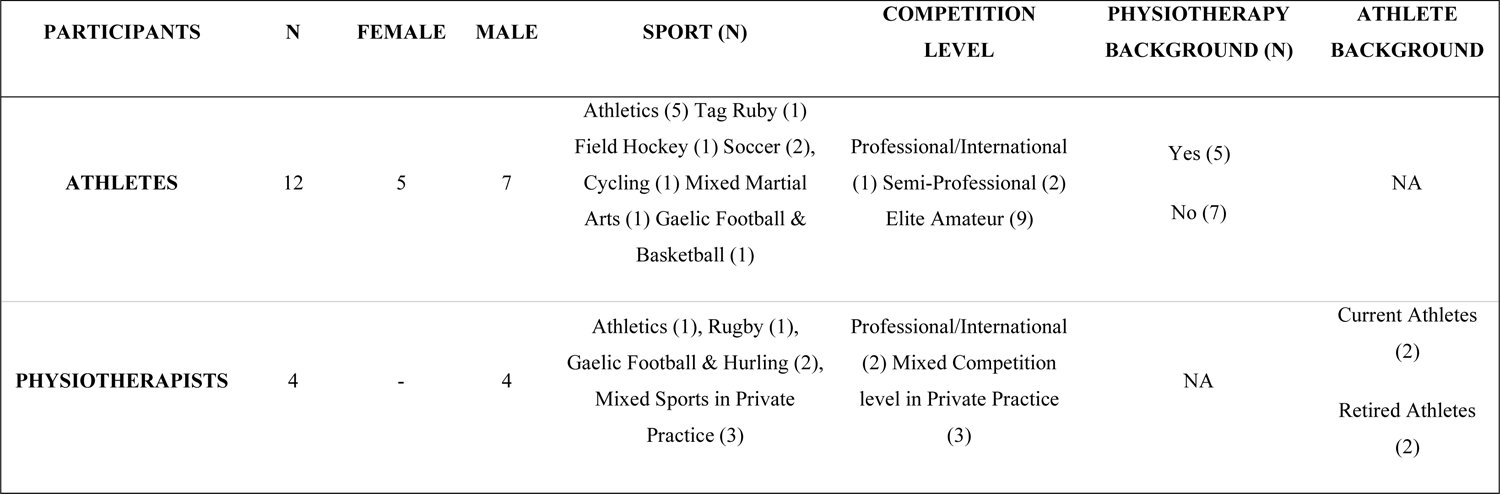
Participant Demographics.

| **PARTICIPANTS** | **N** | **FEMALE** | **MALE** | **SPORT**<br>**(N)** | **COMPETITION**<br>**LEVEL** | **PHYSIOTHERAPY**<br>**BACKGROUND**<br>**(N)** | **ATHLETE**<br>**BACKGROU**<br>**ND** |
| --- | --- | --- | --- | --- | --- | --- | --- |
| **ATHLETES** | 12 | 5 | 7 | Athletics (5) Tag<br>Ruby (1) Field<br>Hockey (1) Soccer<br>(2), Cycling (1)<br>Mixed Martial Arts<br>(1) Gaelic Football<br>& Basketball (1) | Professional/Inter<br>national (1) Semi-Professional (2)<br>Elite Amateur (9) | Yes (5)<br>No (7) | NA |
| **PHYSIOTHERAPIS**<br>**TS** | 4 | - | 4 | Athletics (1),<br>Rugby (1), Gaelic<br>Football & Hurling<br>(2), Mixed Sports<br>in Private Practice<br>(3) | Professional/Inter<br>national (2)<br>Mixed<br>Competition level<br>in Private Practice<br>(3) | NA | Current<br>Athletes (2)<br>Retired<br>Athletes (2) |

| PARTICIPANTS | N | FEMALE | MALE | SPORT (N) | COMPETITION<br>LEVEL | PHYSIOTHERAPY<br>BACKGROUND (N) | ATHLETE<br>BACKGROUND |
| --- | --- | --- | --- | --- | --- | --- | --- |
| ATHLETES | 12 | 5 | 7 | Athletics (5) Tag Ruby (1)<br>Field Hockey (1) Soccer (2),<br>Cycling (1) Mixed Martial<br>Arts (1) Gaelic Football &<br>Basketball (1) | Professional/International<br>(1) Semi-Professional (2)<br>Elite Amateur (9) | Yes (5)<br>No (7) | NA |
| PHYSIOTHERAPISTS | 4 | - | 4 | Athletics (1), Rugby (1),<br>Gaelic Football & Hurling (2),<br>Mixed Sports in Private<br>Practice (3) | Professional/International<br>(2) Mixed Competition<br>level in Private Practice<br>(3) | NA | Current Athletes<br>(2)<br>Retired Athletes<br>(2) |

### Theme 1. Measures Scales and Dimensions

Theme 1 includes three subthemes; 1.1 – value and limitations of tools and scales, 1.2 – multidimensional methods, and 1.3 – making sense and interpreting. Despite positive advances in identifying and managing sports-related pain the nebulous nature of pain experience proves challenging when it comes to accurate and comprehensive assessment.^3^ Participants in this study contrasted the value of tangible measures and scales which add a level of objectivity to the subjective experience of pain with the limitations of currently available tools and scales for assessing athlete pain. Additionally, the need for wider biopsychosocial and multidimensional pain assessment strategies is firmly established in the literature, something with which participants concurred but rarely encountered.^2^ ^4^

### 1.1 Value and limitations of tools and scales

Participants discussed aspects of a standard physiotherapy pain assessment when recalling previous experiences, including observation of the athlete and measures of physical function such as range of motion, strength and performance markers. Physiotherapists and athletes alike noted the importance of pain provocation and localisation to identify the “athlete’s pain” to validate the athlete’s specific pain experience which is promoted by contemporary pain standards.^15^ Physiotherapists acknowledged how criteria-based rehabilitation and pain management, guided by objective indicators can provide clear goals for athletes and preferred this strategy to focusing on timelines which can be difficult to predict. Additionally, athletes appreciated having tangible measures to both represent their pain and gauge progress, a concept that is firmly established in sports-related injury and can also be applied to the assessment of pain.^16^

> “Mobility, and kind of, my range..strength on both legs, that would have been tested, and then pinpointing the pain, physically.” – A11

> “I do a lot of like single leg hopping and stuff like that as I often say to a runner if you can’t single leg hop, you can’t run, you kind of set your parameters early.” – P01

> “ I appreciated that like there was an objective outcome.. (Physiotherapist) being able to see if pain is actually affecting me. There was something that was tangible…I was quite happy that meant he was going to take my pain serious(ly).” – A08

However, in cases where pain was more diffuse or difficult to pinpoint, the limitations of representing pain in a clinical setting were a source of frustration for athletes. Additionally, some athletes experienced negative consequences from pain provocation testing or repeatedly being asked about symptoms, highlighting the need for the selective use of certain pain assessment tools. Choosing the right time to assess pain response and ask about symptoms is an important aspect that has been previously discussed by established pain clinician-researchers.^17^

> “I couldn’t tell where it was I couldn’t point to it, I couldn’t palpate it they couldn’t locate where it was.. which kind of frustrated me.” – A03

> “When I single leg hopped I then had seven out of ten pain for the next two days when I could have told them that ‘this is going to be really sore’.” – A03

The development of technology in sport science and medicine has facilitated additional means for capturing data that can be helpful in the assessment of pain in athletes including wearable devices for physical performance and physiologic data, as well as smartphones and applications for assisting with subjective health and wellness monitoring.^18^ Athletes reported familiarity with physiological markers such as heart rate metrics however, effective use requires planning, implementation and reporting and at times athletes felt overwhelmed with the amount of information.

> “I’m like a real techy person like I’m always on Garmin and Whoop and Auras and all the different things…I went back to physios, went back to Doctors, because my resting heart rate would have been normally you know low forties.. and then for a good two months, it was mid-sixties.” – A10

> “Maybe it was a bad thing that I was keeping such a close eye on you know the data I was collecting yes because I was probably forming some sort of psychological response.”– A10

Questionnaires and pain diaries can be enlisted as a method to add a layer of objectivity to the retrospective subjective reporting of pain.^19^ Athletes, describing the individual nature of preferences for communicating pain, noted how written diaries allow pain to be reported at multiple time points throughout the day addressing the limitations of the once-off point-in-time assessments.

Physiotherapists also noted that written methods add an opportunity for athlete reflection. Conversely, the time taken to complete them was seen as burdensome. Athletes felt they were repeating what had already been expressed through conversation, and physiotherapists acknowledged that they are often not appropriately analysed and utilised, citing time restrictions, and instead placed higher value on objective measures.

> “..where you kind of pull out a sheet and describe the pain and it kind of gets the flow of thought going and you’re better able to communicate it then to the physio.’’ – A11

> “Sometimes there’s a period of time where like pain diaries and you know getting patients to write down every day what the pain was and what they were doing.” – P01

> “ A pain diary.., that takes persistence that I didn’t have.” –A08

Various pain scales are used to gauge the severity of pain an athlete feels during the clinical encounter including during pain provocation tests, at specific time points throughout the day, and following certain activities.^20^ The numerical pain rating scale (NPRS), or 0-10 pain scale, was most frequently encountered by participants who found it may be effective for repeated use with an athlete particularly when tracking large changes in pain over time. However, both athletes and physiotherapists advised comparisons between athletes should not be made. The benefits (simplicity, ability to track large changes in pain in an athlete and widespread use) and drawbacks (once-off measure not representing the 24-hour pain experience, floor/ceiling effects, subjective interpretation of scale) of the NPRS described by participants are closely aligned with established pain measurement literature.^20^

> “I often find with the 0-to-10 scale, someone can say a 5, doesn’t really mean a whole lot. If someone says a 9, it probably does, but if someone says a 3 or 4 that could be very different to someone else’s 3 or 4,., I think it certainly gives me an insight into just that person, and if your pain goes from a 6 to a 2, I know that that’s an improvement for you.” – P04

> “So usually I get asked to describe it from nought to ten but I don’t really get the opportunity to describe the twenty-four-hour cycle of the pain, which I think is probably something maybe more relevant …” – A08

Athletes often struggled to represent their pain as a number and preferred descriptions that they could attribute greater meaning to. Physiotherapists also used alternative pain scales offering different means of measuring and interpreting pain severity including the traffic light system which indicates when athletes must stop activities (red), proceed with caution (amber) or continue unabated (green), an option that has been adopted widely in managing tendon pain.^21^ Grading pain as mild, moderate or severe was another option participants encountered.

> “I find it very hard to say nought to ten so she would say mild, moderate or severe on the pain scale like something that took it away from me giving it a number.” – A08

### 1.2 Multidimensional methods

Athletes emphasised the importance of accurately representing the psychological/emotional aspects (e.g. feelings and emotions related to pain and sport), wider biological and lifestyle factors (e.g. stress, sleep and nutrition, age and time of a female athlete’s menstrual cycle), and environmental factors (e.g. pain culture within the sport, time of the competitive season, weather and playing conditions).

Athletes highlighted the profound impact these aspects have on pain perception, yet despite best practice guidance, were rarely encountered.^2^ ^4^ Physiotherapists, acknowledging that their inclusion is best practice, shared that some formal assessments of psychological and emotional aspects can be lengthy and interrupt the flow of assessment in time-pressured sports settings where athletes may not appreciate them. Instead, physiotherapists described how these aspects tend to be addressed more informally as part of the pain interview. Techniques such as motivational interviewing and psychologically informed practice are becoming standard practice for physiotherapists in the general population and likely inform the assessment process for physiotherapists working in sports also.^22^

It is worth highlighting how athletes in this study felt these aspects were often ignored and so there may be a case for explicitly discussing these wider aspects to ensure athlete’s pain experience is validated.

> “I know personally if my cortisol levels are higher, so if I’m stressed or lacking sleep that I nearly feel pain more and as a woman actually at different parts of your cycle.” – A08

> “I think people probably didn’t ask enough about the mental effects of injury until you are visibly in a bad mood or something” – A05

> “On the psychological stuff, I suppose the way I would normally do it is you kind of a little more informal so you kind of be having a chat with them.” – P02

### 1.3 Making sense and interpreting

Prior research has noted how physiotherapists can sometimes feel underequipped when assessing pain, particularly with more complex and chronic presentations which has led to the recent publication of pain education curricula at undergraduate and postgraduate levels by the European Pain Federation.^22^ ^23^ Drawing on their experience of prior pain assessments, athletes highlighted the need to go beyond the available pain scales and search for better ways to describe and represent pain intensity. Physiotherapists also acknowledged how in their experience objective findings must be correlated with athletes’ subjective reports of pain to complete a comprehensive pain assessment that is in line with best practice guidance and the recent pain education curricula.^2^ ^23^

> “Instead of like tell me what your pain is from nought to ten it’s like tell me how it impacted your ability to train, your ability to race… to live a normal life.” – A08

> “The outcome measures.. like handheld dynamometry and force plates.. you’re trying to correlate that with their own.. feedback on pain.” –-P01

### Theme 2. Connect, Listen & Learn

Theme 2 includes two subthemes; 2.1 – the pain interview and athlete’s story and 2.2 – forging the athlete-clinician connection. Every athlete has their own unique “pain story” and effectively facilitating an athlete to describe the meaning their pain experiences holds for them is a cornerstone of pain assessment.^17^ This aspect of assessment was seen by athletes and physiotherapists as an opportunity to develop a rapport and relationship with characteristics such as trust, authenticity and empathy woven into the process. A well-conducted pain interview allowed athletes to feel listened to, understood and validated, as prioritised in pain assessment guidelines.^15^ In contrast, assessments in sports settings are often time-pressured with establishing injury diagnosis being prioritised meaning comprehensive pain assessment and optimal management cannot be achieved.^24^

### 2.1 The pain interview and athlete’s story

The pain interview allows the athlete and physiotherapist to explore the sensory and emotional aspects of pain and their wider impact.^2^ Athletes discussed being asked about aggravating and easing factors, severity, irritability and nature of the pain and to describe it using various adjectives. The onset, duration and preceding history of the pain experience were commonly explored. Athletes and physiotherapists emphasized how the pain interview is an interactive process that is enhanced by the physiotherapist exploring the context and history behind an athlete’s pain experience. Athletes gathered pain and injury experiences from their support network and teammates adding them to their interpretations. Physiotherapists used the pain interview to guide the generation of hypotheses and the selection of objective tests and measures.

> “Because everyone has pain, no one goes through life without pain, and then it’s individual to that person so maybe help give them a bit of context to their own pain; this feels.. similar to what I had before,.. to what this person told me they had, or… “ – P04

> “If we don’t have an idea before they’re on.. the treatment table,.. of two or three things that might be going on here well then we either haven’t asked the right question or we haven’t listened to them.” *–*P01

Open-ended discussions allowed the athlete to go beyond the sensory or physical manifestations of pain and begin to explore the emotional and multidimensional aspects. Open-ended questions aided the clinician in gathering the necessary information needed to develop accurate hypotheses and diagnoses in line with research findings of question styles used in other pain cohorts.^25^ Conversely participants found the judicious use of closed questions were helpful to focus on specific aspects of the pain experience.

> “Just letting someone talk to me about what their pain is, you learn a bit about the physical side of it but then, you know, what does their pain means to them.. the emotion side of it and the mental side of it as well, you get a bit more information out of it..,,.. a lot of the time people will tell you exactly what’s wrong with them” –P04

### 2.2 Forging the athlete-clinician connection

Athletes valued having an opportunity to give their unique and full perspective which lay the foundation for a strong relationship with their physiotherapist. Listening to an athlete with empathy provides validation and reassurance that they are being heard and understood enabling effective therapeutic alliance and an effective working relationship.^15^ Participants shared how truly understanding an athlete’s pain experience requires an investment of time and energy.

> “I think it really matters how you talk to someone about their own pain you know if they think that it’s really important to them, it’s really important.” – A03

> “This is the first time someone’s actually listened to what’s been going on and like. I think.. you tend to get a little more insight into their pain.” – P01

Athletes and physiotherapists alike stressed the importance of seeing the athlete as a whole person, prioritising human interaction and connection and understanding their emotions, motivations and behaviours. Trust plays an integral role in fostering the athlete-physiotherapist relationship and an athlete’s confidence in the diagnosis, pain explanation and prognosis they have received.^26^ The ambiguous nature of pain and the pressure to decide on whether or not to play and/or train through pain in high-stakes sports settings can create barriers around forming strong relationships. This may be due to sports physiotherapists’ disclosure obligations to the management team or due to the unwillingness of athletes to disclose the full extent of their pain. The athlete pain assessment was described as a delicate balance between capturing sufficient objective data to structure the assessment and management and dedicating time to nurture relationships and forge connections.

> “I think it’s all just about understanding people really and then obviously you have to have the rationale to back up what you’re going to do etc..” – P02

> ‘’There’s a balance I suppose in terms of seeing people as a number on a sheet and also as a human being. Sometimes.. with outcome measures. It ends up where people just don’t be seen at all as a person.’’ – P03

Both athletes and physiotherapists being open to different methods of communication, assessment and management is an important aspect previously highlighted in the literature that participants resonated with and felt this helped achieve an effectively balanced and individualised assessment.^27^

> “Understanding how people communicate is good because some people are, you know, would be more comfortable writing, some people are comfortable with verbal, some people might be more comfortable showing, you know..” – P04

> ‘In an ideal world it’s like every patient gets a very individual service I suppose… the person, their previous experiences, you know their goals, their objectives this session like you know so then you can (use) different parts of you know assessment tools and then you can go dictate your treatments.’’ – P03

### Theme 3. Lighthouse In The Storm: providing direction for athletes in pain

Theme 3 includes three subthemes; 3.1 – information overload and indecision, 3.2 – a beacon of direction – the role of the physiotherapist and 3.3 – the burden of expectation – challenges for physiotherapists. Understanding and making sense of pain can be challenging for athletes. They found that the interaction with a physiotherapist helped them make informed decisions relating to their pain. The role of providing clarity and direction for athletes is burdensome.^28^ Time demands, high expectations and work environment difficulties can be likened to the solitary and challenging role of the lighthouse keeper. Research has highlighted how physiotherapists can sometimes feel challenged in particular when managing chronic pain.^22^

Whilst it is important to seek support from other members of the interdisciplinary team when appropriate, physiotherapists are core members of the pain team with expertise in exercise prescription and psychologically informed care who are optimally positioned to guide the pain management journey for athletes.

### 3.1 Information overload and indecision

Athletes described their frustration to accurately represent their pain during the assessment process. Additionally, addressing the potential for pain hypervigilance, athletes noted that too much attention amplified pain experiences and ruminating thoughts intruded. In line with previous findings, athletes and physiotherapists noted the assessment of pain can be further complicated by cognitive overload through an abundance of information gleaned from social media, support networks and various clinicians that can leave an athlete feeling confused and frozen in indecision.^29^

> “You kind of have that feeling of, you know, is that pain or is that, you know, your mind, because you’re overthinking when you’re coming back.. that’s something that I definitely struggle with.” – A12

> “Like Instagram and people influencing.. I was changing exercises every single day trying to solve my pains and everything.. the stress and the thought process and then doubting other therapists was another huge thing so like.. there’s a lot of therapists that clash and it just doesn’t help at all.” – A07

### 3.2 A beacon of direction – the role of the physiotherapist

Athletes highlighted how a clear explanation of their pain presentation provides clarity and reassurance helping them to accept their current situation and begin to plan their next steps. The pain assessment is an opportunity to educate athletes about the causes and contributors to their pain perception which can have a positive effect on pain experience and management and sports performance.^3^ ^5^

> “I know exactly what it is, and I nearly come to terms with it an awful lot quicker whereas if it’s an injury that I’m not sure what it is sometimes I feel like my interpretation of that is worse. I feel like my education around it has a huge impact so therefore that knowledge aspect is a drawback if I’m not asked about it”. – A08

> “As a kind of full-circle to the person.. “this is what the pain is” or “this is what I think it is and this is why I think it is” and “this is what we’re going to do about it” and then if you understand the first two then the third becomes a lot easier for the person to actually continue on because they understand, you know” – P04

### 3.3 The burden of expectation – challenges for physiotherapists

Physiotherapists acknowledged the challenging nature of assessing and understanding pain and the difficulty of choosing the appropriate pain assessment tools and measures. During the clinical reasoning process, physiotherapists must be mindful of their communication strategy with athletes. With due regard to the “information overload’’ described by athletes, physiotherapists highlighted how they were conscious not to cause further confusion in the pain assessment process, which can be a challenge.

> “Sometimes you come away from it and you wanted, why didn’t I do that, why did I do this, we should assess this, I didn’t actually. You know, it can be hard.” – P03

Striking the balance between assessment tools and diagnosis is a fine art, when not satisfied with the pain explanation, diagnosis or prognosis provided by one clinician, athletes may seek additional assessment, something clinicians were acutely aware of. Taking a health literacy-sensitive approach is essential to ensure patients can be actively involved in the decision-making about their own care. Health literacy impacts a variety of health outcomes in patients with chronic pain and should also be prioritised with athlete cohorts to ensure optimal management.^30^

> “All I can remember is I can’t run for six weeks so that’s not going to happen so I’ll go to somebody else and find the answer I want” – P01

Physiotherapists also described the challenges of completing pain assessments pitch side or in other sports settings with conditions often less than optimal to conduct a thorough assessment. The timing of the assessment is a key consideration with early and repeated assessments recommended in musculoskeletal physiotherapy standards of practice.^30^ This gives a clearer and more comprehensive picture of pain. In contrast to these standards, the pain assessment practice described by athletes was challenged by the lack of time available in both private practice and sports physiotherapy setting.

Additionally, the pressure to return to performance and minimise time out of sport was highlighted, an aspect that has been previously discussed by sports physiotherapists.^28^ Physiotherapists acknowledged the difficulties of completing a thorough assessment and providing a diagnosis from the initial assessment.

> “Like if you’re working with an athlete day-to-day then it’s a lot easier to gauge pain versus if I have someone that comes in off the street into the clinic that I’ve never met before, and you don’t know their tendencies or you don’t know their history.” – A12

> “I think it’s like, putting the pressure on yourself to know exactly what’s wrong with someone the first time you see them is unrealistic” –P04

## Conclusion

We described and explored the phenomena of pain assessment in sport. Athletes and physiotherapists described and critiqued the routine methods and measures used. We highlighted the value of the pain interview and athlete’s story and we discussed how combining objective findings with the athlete’s pain experience requires consideration and skill. Athletes shared their desire for direction in understanding and managing pain and we highlighted the challenges this poses, particularly for physiotherapists. Whilst every effort was made to collect experiences from a diverse range of athletes and physiotherapists and variety was achieved in sport, competition level and practice setting the experiences gathered from these focus groups may not apply to all athlete pain assessment settings. Notably, participants were all recruited from Ireland and whilst some of the female athletes also had a physiotherapy background, no female sports physiotherapists were available to participate.

Better, more comprehensive and multidimensional means to describe and assess pain are priorities for research and practice. Additionally, improved communication strategies that facilitate more timely and relevant pain information whilst preserving an effective athlete-physiotherapist relationship are needed. Priorities and directions for pain assessment will be explored further in Part Two.

### Practical Implications

– Objective assessment tools offer value in supporting the subjective nature of pain experience and as indicators of progress.
– Some commonly used objective tools and scales have their limitations when it comes to comprehensively assessing pain. The overuse of objective measures that may not be relevant or helpful should be avoided.
– Multidimensional psychosocial assessment tools are underused and Physiotherapists should consider integrating them into their athlete pain assessment practice.
– Physiotherapists must place a high value on a well-conducted pain interview to (i) facilitate the athlete to tell their pain story, (ii) educate the athlete regarding their pain and (iii) as an opportunity to develop the therapeutic relationship.
– Physiotherapists must include multidimensional pain assessment measures and take an individualised approach when interpreting pain assessment findings.

## Data Availability

All data produced are available online at https://data.mendeley.com/datasets/t47tw94mzd/2

