## Appendices for "Exploring athlete pain assessment experiences and priorities; A two-part qualitative series of athlete and physiotherapist interactions. Part One. “Gauging and discerning” - Athlete & physiotherapist pain assessment experiences and interactions"

**Appendix A Focus Group Topic Guide/Question Route**

*First 10-15 mins. Ground rules (equal voices, respect opinions confidential nature of session) Participants are invited to introduce themselves, say a little about themselves*

**Question 1) What does pain mean to you in terms of being an athlete?**

Prompts: Are there different types of pain? Injury vs training soreness? Do you think pain means different things to different athletes? Is short-term pain different to long-term pain?

**Question 2) Can you recall an upper or lower limb pain experience you had in the last year?**

Prompts: Was it an acute pain episode or recurrent/lingering? What were the physical feelings/sensations associated with that pain? What were the emotions and thoughts associated with that pain? When was it, what time of the year/season? How did it change your day-to-day and sporting activities? How did you manage/address this pain?

**Question 3) Can you recall how your upper or lower limb pain was assessed?**

Prompts:/Probes: What did the assessment involve? Did they ask questions or discuss different aspects of pain? Did they carry out any tests or measurements? Where did the assessments take place? Was anyone else involved in the assessment process? Was there a difference in the different people you interacted with and how they approached pain eg coaches, doctors/surgeons, mental health professionals etc

**Questions 4) What are some of the drawbacks of pain assessments you have experienced or encountered?**

Prompts: These may include specific questions, tests, tools, or ideas that you may have encountered

**Questions 5) What are some ways pain assessments could be improved in the future? Can you think of any interesting or innovative ways or methods to evaluate pain?**

Prompts: These may include specific questions, tests, tools, or ideas that you may have encountered

**Appendix B** **The Pain Assessment Priority Pyramid**.

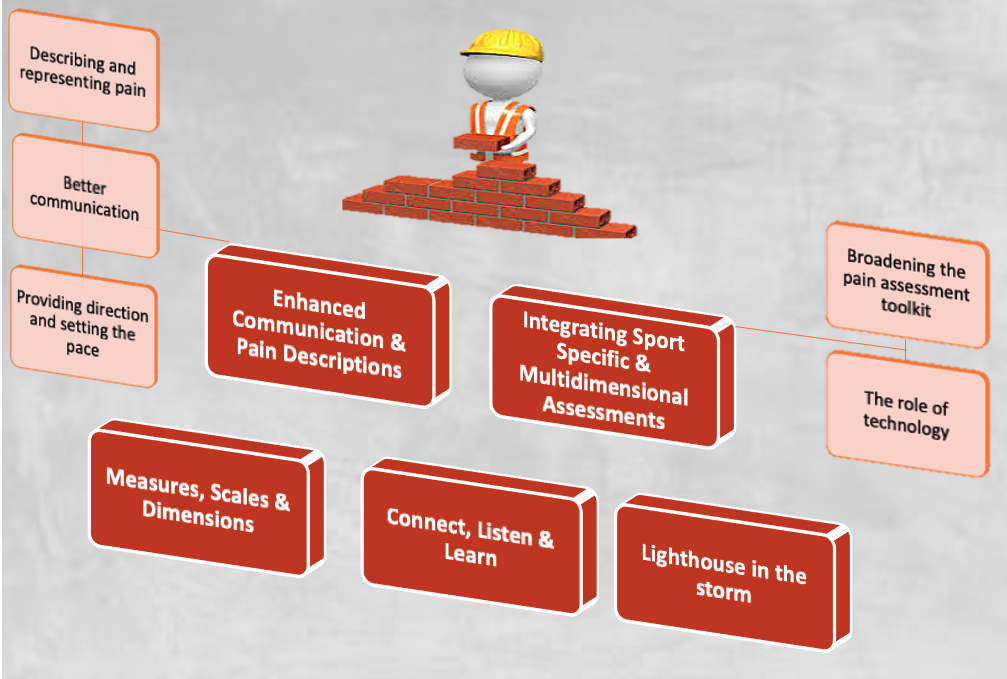

This pyramid displays the 5 themes developed over the course of the focus groups. The bottom row or foundation of the pyramid comprises the three themes that describe the “athlete pain assessment” experience. The top row, the “priorities and directions for athlete pain assessment” themes and subthemes that will be presented in Part Two of this series therefore “build” on the themes from Part One.

Source: The cartoon element at the top of this image was designed by Freepik [www.freepik.com](http://www.freepik.com/)
